# First records of *Triatoma sanguisuga* (Hemiptera: Reduviidae) and detection of *Trypanosoma cruzi* (Trypanosomatida:Trypanosomatidae) in southern New Jersey, United States

**DOI:** 10.64898/2026.10.27.26366644

**Authors:** Heidi E. Herb, Alexander R. Kelley, Jennifer K. Peterson, Dina M. Fonseca

## Abstract

*Triatoma sanguisuga* (LeConte, 1855) is the most widely distributed triatomine bug species in the United States (US), but its occurrence and association with *Trypanosoma cruzi* (Chagas, 1909) are poorly characterized near the northeastern margin of its reported range. We combined homeowner-submitted specimens, community-science records, targeted habitat surveys, opportunistic light-trap collections, molecular identification, and pathogen screening to characterize *T. sanguisuga* and *Trypanosoma cruzi* in New Jersey. Archived mammal blood samples from central New Jersey were also screened for *Trypanosoma cruzi*. *Triatoma sanguisuga* was documented at multiple locations in southern New Jersey, including nymphs collected from standing dead trees and stumps and adults attracted to light traps. Mitochondrial *cox1* sequences from New Jersey specimens clustered with sequences from *T. sanguisuga* collected in Delaware and suggest geographic structure among US populations. *Trypanosoma cruzi* DNA was detected in adult but not nymphal triatomines from southern New Jersey; parasite DNA was not detected in archived mammal samples from central New Jersey. These findings provide the first verified field collections of *T. sanguisuga* in New Jersey and the first detection of *Trypanosoma cruzi* in triatomines from the state. Although the opportunistic sampling design does not permit estimates of statewide distribution, abundance, or infection prevalence, the findings establish an initial baseline for investigating triatomine distribution, parasite occurrence, and population structure at the northeastern extent of the species’ range.

## Introduction

The geographic distribution and abundance of arthropod vectors are fundamental determinants of vector-borne disease risk, but these patterns are often poorly resolved near the margins of species’ known ranges, particularly where systematic surveillance is limited (Bender et al. 2020). Triatomine bugs (Hemiptera: Reduviidae), which can vector the causative agent of Chagas disease, are most diverse in tropical and subtropical regions, but several species extend into temperate North America. Near the margins of their known ranges, occurrence records can be sparse, historical, or lack precise locality information, making them difficult to confirm.

Resolving these distributional gaps is important because identifying where competent vectors and their associated pathogens occur informs estimates of transmission risk and the targeting of public health interventions (Gutfraind et al. 2018). Knowledge of vector and pathogen distributions may also influence healthcare provider awareness and, consequently, the timeliness of diagnosis and treatment (Olivera et al. 2018, Adams et al. 2026).

Chagas disease is caused by the kinetoplastid protozoa *Trypanosoma cruzi* (Chagas, 1909), which is transmitted when excreta from *Trypanosoma cruzi-*infected triatomine bugs are deposited near open wounds or mucous membranes (Eickhoff et al. 2013). Infection with *Trypanosoma cruzi* in humans can be chronic and, if left untreated, can lead to serious cardiac or gastrointestinal complications (Bern et al. 2011). Recent estimates put global Chagas disease prevalence at around 10 million people (Condray and Newman 2026). There are no vaccines for Chagas disease and treatment is effective only during the initial stage of infection (Bern et al. 2011).

*Triatoma sanguisuga* (LeConte, 1855) is the most widely distributed of the 11 triatomine species known in the United States, with a reported range spanning much of the eastern and central United States from the Rocky Mountains to the Atlantic seaboard (Bern et al. 2011, Reeves and Miller 2020, Peterson et al. 2024). Most ecological and epidemiological studies of *T. sanguisuga* have focused on populations in the southern United States, particularly in Texas, Louisiana, and Florida, where the species has been found infected with *Trypanosoma cruzi* and has been documented entering human dwellings and feeding on a broad range of vertebrate hosts, including humans (Kjos et al. 2013, Waleckx et al. 2014, Beatty et al. 2025). In contrast, comparatively little is known about *T. sanguisuga* near the northern extent of its range, particularly in the northeastern United States (Peterson et al. 2024, Santos et al. 2024).

New Jersey lies near the northeastern limit of the reported range of *T. sanguisuga*, yet the species has not been formally studied in the state. Although New Jersey has been included in published range summaries (Bern et al. 2011) we found no associated locality data or documented voucher specimens that would allow those reports to be independently verified.

More direct evidence emerged in 2016 and 2023, when specimens subsequently identified as *T. sanguisuga* were submitted by New Jersey residents to Rutgers (Joseph Ingerson-Mahar, Vegetable IPM Coordinator, and Salvatore Mangiafico, Salem County Extension Agent, *personal communications*). In addition, photographs identified as of *T. sanguisuga* have been submitted to iNaturalist since 2018 (GBIF.org 2026). Collectively, these records provide compelling evidence that *T. sanguisuga* occurs in New Jersey, but its distribution, abundance, and *Trypanosoma cruzi* infection prevalence remain unresolved.

Given the public health importance of triatomine bugs as vectors of *Trypanosoma cruzi*, we sought to reduce key uncertainties regarding the occurrence, distribution, and infection status of *T. sanguisuga* in New Jersey. Because resources were insufficient for a systematic statewide survey, we combined multiple complementary data sources, including published records, community-science observations, the two homeowner-submitted specimens received earlier, and targeted field surveys. We also performed *Trypanosoma cruzi* screening on available and newly collected triatomine specimens and archived mammal samples. Our objective was to establish a preliminary eco-epidemiological baseline for *T. sanguisuga* and *Trypanosoma cruzi* in New Jersey to identify priorities for future surveillance.

## Materials and Methods

### Archived specimens

Two adult female triatomines were submitted by New Jersey (NJ) residents, one from Cumberland County in June 2016 and another from Salem County in August 2023. The 2016 specimen was initially submitted to the Rutgers University Vegetable Integrated Pest Management Program, and the 2023 specimen was submitted to the Rutgers Cooperative Extension office in Salem County. According to the Rutgers personnel who received and identified the specimens, both were found by homeowners in their bedrooms (Joseph Ingerson-Mahar, Vegetable IPM Coordinator, and Salvatore Mangiafico, Salem County Extension

Agent, *personal communications*). We obtained the Cumberland specimen in 2019 and the Salem specimen in 2023. After receipt, the specimens were kept dry and stored at room temperature until further processing. They have since been pinned, labeled, and submitted to the Rutgers Entomology Collection (Accession numbers TBD).

### Field surveys

#### Habitat surveys

On 26 February 2025, we conducted a field survey at the Thundergut Pond Wildlife Management Area, Salem County, NJ (GPS coordinates 39.542933,-75.312328). This location was selected due to its proximity to the collection location for the archived specimen from Salem

County. Common habitat types include deciduous and mixed upland and wetland forest (NJ Department of Environmental Protection n.d.). On 12 March 2025, we conducted a second field survey on Rutgers University Cook Campus in New Brunswick, NJ, located in Middlesex County. Two forested sites were searched: “Goat Farm” (40°28′22.9″N, 74°26′14.2″W), a forested patch adjacent to a grassy area and goat and sheep pens, and “University Inn” (40°29′02.2″N, 74°25′48.7″W), a forested patch adjacent to a meadowed area and the Rutgers University Inn & Conference Center. Both sites were predominantly deciduous forest, including oak and maple trees (Breden et al. 2001). At all sites, we searched standing dead trees in forested areas for triatomines by removing pieces of bark and inspecting the interior of the bark, the exterior of the exposed trunk, and any accessible insect cavities for *T. sanguisuga*.

All specimens resembling triatomines were placed in collection jars with cardboard pieces for insect shelter and were transported alive to the Rutgers Center for Vector Biology (CVB) in New Brunswick for further processing. Adult triatomines, including archived and newly collected specimens, were sexed using external morphological characteristics following Beatty et al. (2023). We identified non-triatomine specimens using appropriate taxonomic keys (Hoebeke et al. 2019). Collections were carried out under a Special Use Permit from the NJ Department of Environmental Protection (issued 13 December 2024).

#### Light trapping

Triatomines were collected opportunistically at light traps operated by a local naturalist at a private residential property in Cape May County, New Jersey, from 2022 to 2025. The traps were deployed for moth and other nocturnal insect observations. Two light arrays were used on the property, including a mercury vapor/LED blacklight setup near a shed and an Entoquip LED blacklight setup along the property fence line. Lights were operated overnight on automatic timers, and the arrays were checked periodically during nighttime observation periods. Beginning in 2025, triatomines were kept alive in plastic containers with holes until they were picked up and taken to Rutgers CVB for processing.

### DNA extraction

DNA extraction was performed on all insect specimens using DNeasy Blood and

Tissue spin-column kits (Qiagen, Inc. Germantown, MD, USA). For each specimen, we made an incision into the abdomen and removed material from the gut for DNA extraction. For adult triatomine specimens, we also separately extracted DNA from a leg to use as a control for pathogen detection. We followed the manufacturer’s protocol with minor modifications to the final elution step, where we eluted in 75μL of buffer AE to increase DNA concentration. For archived specimens, we performed additional DNA extractions with a final elution in 50μL of buffer AE to further increase DNA concentration.

### DNA barcoding

To confirm species identification for insect specimens, we used a standard DNA barcoding PCR protocol using the primers HCO2198 (5’-TAAACTTCAGGGTGACCAAAAAATCA-3’) and LCO1498 (5’-GGTCAACAAATCATAAAGATATTGG-3’) to amplify a 710 base pair (bp) region of the cytochrome oxidase subunit I (*cox1*) gene (Folmer et al. 1994). Thermocycling conditions consisted of a touchdown protocol including 5 cycles of 96°C for 30 seconds, 53°C to 48°C (decreasing 1°C for each subsequent cycle after the first) for 30 seconds, and 72°C for 45 seconds, followed by 40 cycles of 96°C for 30 seconds, 49°C for 30 seconds, and 72°C for 45 seconds, with a final extension at 72°C for 10 minutes. We used gel electrophoresis on a 1% agarose gel to visualize PCR products and amplicons of the expected size were cleaned with ExoSAP-IT (Thermo Fisher, Waltham, MA, US) using the manufacturer’s protocol. Cleaned PCR products were sent to Azenta Genewiz (South Plainfield, NJ, US) for unidirectional (non-triatomine specimens) or bidirectional sequencing (triatomine specimens). For sequences obtained with bidirectional sequencing, we trimmed and aligned obtained sequences using Geneious Prime 2025.0.3 (Biomatters Inc., San Diego, CA, US) to generate consensus sequences.

### Mammal samples

Archived DNA extracted from mammal blood samples from a previous trapping study conducted in Middlesex County, New Jersey (Ferreira et al. 2023, Herb et al. 2026) was available for *Trypanosoma cruzi* screening. Samples were originally collected from small-to medium-sized mammals on the Rutgers University Cook Campus between May and September 2021 and stored at −80°C until testing. Mammal trapping locations included the “Goat Farm” and “University Inn” locations where we performed the field surveys for triatomines in March 2025. We prioritized screening mammal species that were previously documented as positive for *Trypanosoma cruzi* or as blood-meal hosts of *T. sanguisuga* (Hodo & Hamer, 2017). In total, 138 blood samples representing 126 individual mammals from seven species were available for screening (Table 1). Animal capture and sampling procedures were conducted under Rutgers University IACUC protocol PROTO201900163 and New Jersey Division of Fish and Wildlife permit SC 2021-01.

**Table 1.** Mammal blood samples from Rutgers University Cook campus, May to September 2021, screened for *Trypanosoma cruzi*.

| Mammal Species | Total Samples<br>n (unique<br>individuals)* |
| --- | --- |
| Eastern chipmunk<br>*Tamias striatus* | 36 (34) |
| Eastern cottontail<br>*Sylvilagus floridanus* | 1 (1) |
| Gray squirrel<br>*Sciurus carolinensis* | 36 (34) |
| Groundhog<br>*Marmota monax* | 9 (7) |
| Raccoon<br>*Procyon lotor* | 33 (28) |
| Striped skunk<br>*Mephitis mephitis* | 12 (12) |
| Virginia opossum<br>*Didelphis virginiana* | 11 (10) |
| Total | 138 (126) |

\* For some species, the number of total samples per mammal is greater than the number of unique individuals examined because some individuals were recaptured.

### Pathogen screening

#### Light microscopy

When live triatomine specimens were available, we searched for live trypanosomes in their feces using light microscopy. While restraining the insect by grasping a foreleg with forceps, we applied firm pressure to the caudal abdomen until clear to brown excrement was expressed on a glass slide. We mixed the excreta with approximately 5 µL of phosphate-buffered saline and placed a coverslip over the mixture. To visualize trypanosomes, we observed the slide with a Leica microscope (Model DM750) using the 40X objective.

#### Testing for parasite DNA

To detect *Trypanosoma cruzi* DNA with high sensitivity, we screened mammal and triatomine DNA extractions with a previously published TaqMan probe-based qPCR protocol targeting satellite DNA using primers Cruzi 1 (5’ – ASTCGGCTGATCGTTTTCGA – 3’), Cruzi 2 (5’ – AATTCCTCCAAGCAGCGGATA – 3’) and probe Cruzi 3 (5’ – Fam-CACACACTGGACACCAA-NFQ-MGB – 3’) (Duffy et al. 2013). We used an Applied Biosystems 7500 real-time PCR system (Thermo Fisher, Waltham, MA) with thermocycling conditions of 95°C for 10 minutes followed by 40 cycles of 95°C for 15 seconds and 58°C for 1 minute. Positive controls of *Trypanosoma cruzi* positive triatomines or mammals and negative controls of nuclease-free water were run with each experiment. Due to limited sample availability, we did single reactions for each mammal blood sample. For the triatomine screening, we screened DNA extractions from each bug’s gut in triplicate. Given that *Trypanosoma cruzi* is present in feces of infected bugs and that the bugs collected from light traps were housed together prior to transport to the laboratory, we also screened leg extractions in duplicate as a control for environmental contamination. For one archived triatomine specimen with a high cycle threshold (Ct) value on initial screening, we repeated the qPCR to verify evidence of infection with a more concentrated gut DNA extraction. Due to the potential for DNA degradation during storage, we set different positivity thresholds for archived and fresh field specimens. We considered archived specimens with gut Ct values of <35 and field specimens with gut Ct values of <33 to be positive for *Trypanosoma cruzi* DNA (Curtis-Robles et al. 2018, Beatty et al. 2025). Triatomines with negative gut extractions were considered negative for *Trypanosoma cruzi* DNA regardless of the leg extraction results.

### Phylogenetic analysis

Consensus *cox1* barcode sequences for each specimen were submitted as queries to NCBI’s Basic Local Alignment Search Tool (BLAST) using the GenBank core nucleotide database (core nt) (Camacho et al. 2009). We aligned distinct *cox1* sequences obtained from this study to sequences with the highest pairwise identities as well as sequences from triatomine species known to occur in the US (Bern et al. 2011) available on GenBank. *Panstrongylus rufotuberculatus* (GenBank MF614953) was used as an outgroup. We trimmed the alignment to 581 base pairs and a maximum likelihood tree with 1000 ultrafast bootstrap replicates was constructed using the IQ-TREE web server and ModelFinder (Trifinopoulos et al. 2016, Kalyaanamoorthy et al. 2017, Hoang et al. 2018).

## Results

### Triatomine collection

In February 2025, we collected 5 *T. sanguisuga* nymphs from 2 standing dead trees (‘snags’) and one stump in the Thundergut Pond Wildlife Management Area, Salem County. Three nymphs were found in one snag, one nymph was found in the other snag, and one nymph was found in the stump. In March 2025, no triatomines were found at the sites surveyed in Middlesex County, but one flat bug (Heteroptera: Aradidae), morphologically identified as *Mezira subsetosa* (Hoebeke et al. 2019) was collected at the “Goat Farm” site. From June to September 2025, 22 *T. sanguisuga* adults (12 females and 10 males) were collected from light traps in Cape May. Geographic locations of collections and sampling efforts are detailed in Figure 1.

**Figure 1.**
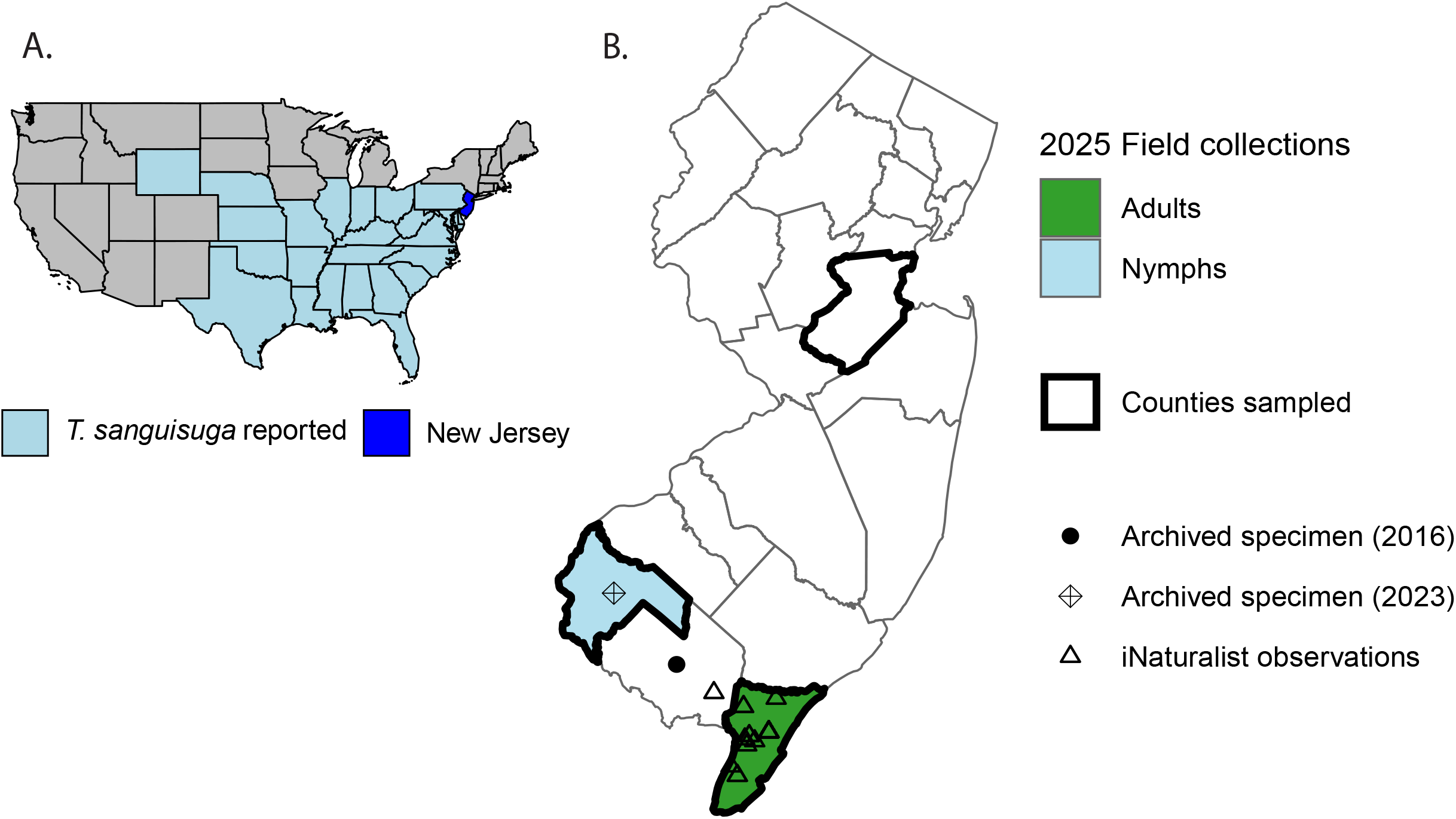
*Triatoma sanguisuga* distribution in the United States and New Jersey, 2026. A. States *Triatoma sanguisuga* has been reported in as of 2026 (Reeves and Miller 2020, Nielsen et al. 2021, CDC 2026). B. *Triatoma sanguisuga* were only found in the southernmost counties of New Jersey despite sampling efforts in northern counties. Archived specimen symbols are placed in the centroid of the counties for confidentiality purposes. iNaturalist observations from 2018– 2026 (n = 55) (GBIF.org 2026). Figure 1 Alt Text Maps indicating where *Triatoma sanguisuga* has been reported, including a state-level map of contiguous United States and specimen collection locations by county in New Jersey.

### Barcoding

Sanger sequencing yielded sequences of 526–630 bp from the 2023 archived specimen, three nymphs, and five field-collected adults (Genbank accession numbers TBD). The archived specimen was only successfully sequenced unidirectionally (630 bp) while the remaining sequences were consensus sequences from bidirectional sequencing (526–621 bp). All triatomine sequences had a top pairwise identity (96.6–97.2%) to a *Triatoma sanguisuga* sequence from Florida (GenBank MT556653) (Aguilera-Uribe et al. 2020). Triatomine sequences obtained in this study were identical with the exception of the 2023 archived specimen and one of the nymphs, which were identical to each other and had one bp difference from the other sequences (99.8% pairwise identity). The sequence obtained from the flat bug specimen had a top pairwise identity (85.9%) to an Aradidae sp. sequence (GenBank OM323175).

### Phylogenetic analysis

*Triatoma sanguisuga* sequences obtained in this study clustered with other *T. sanguisuga* sequences from the northeast, southern, and midwestern US (Figure 2). Within the *T. sanguisuga* clade, both sequences from NJ triatomines closely clustered with a sequence from a *T. sanguisuga* specimen collected in Delaware, while sequences obtained from the midwestern US portion of *T. sanguisuga*’s range formed a separate cluster. The *T. sanguisuga* clade was distinct from all other triatomine species included in the phylogenetic analysis.

**Figure 2.**
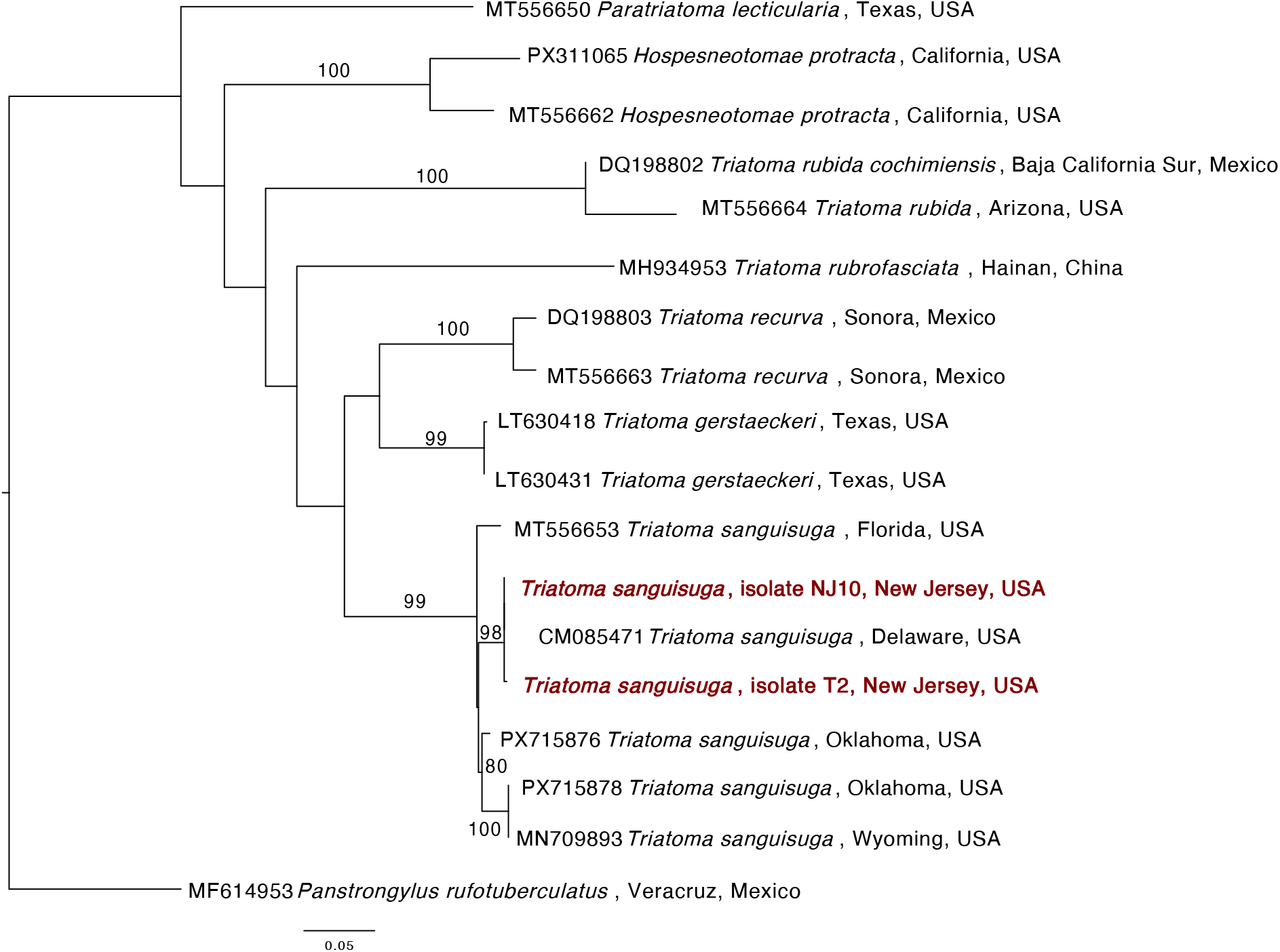
Maximum likelihood phylogenetic tree of triatomine *cox1* sequences, including triatomines found in New Jersey, US, 2025. Sequences from this study shown in red and bootstrap values greater than or equal to 75 are displayed. Phylogenetic tree constructed with TIM2+F+I+G4 substitution model. Figure 2 Alt Text Phylogenetic tree displaying the relationships between *Triatoma sanguisuga* sequences generated in this study and triatomine sequences from the United States and other countries.

### Pathogen detection

Feces from 5 field collected nymphs and 3 adults were examined visually for trypanosomes, and none were observed. *Trypanosoma cruzi* DNA was detected in adult triatomines (14/24, 58.3%), including archived and field-collected specimens (Table 2). Among adults, detection rates were 50.0% (7/14) in females and 70.0% (7/10) for males. None of the 138 screened mammal blood samples nor the 5 triatomine nymphs were positive for *Trypanosoma cruzi* DNA. Of the 14 triatomines that were gut-positive for *Trypanosoma cruzi* DNA, 7 adults were gut-positive and leg-negative while 7 were positive in both gut and leg. Of the 10 triatomines for which we did not detect *Trypanosoma cruzi* in DNA from the guts, 9 were negative in both gut and leg while 1 specimen was leg-positive but gut-negative.

**Table 2.**
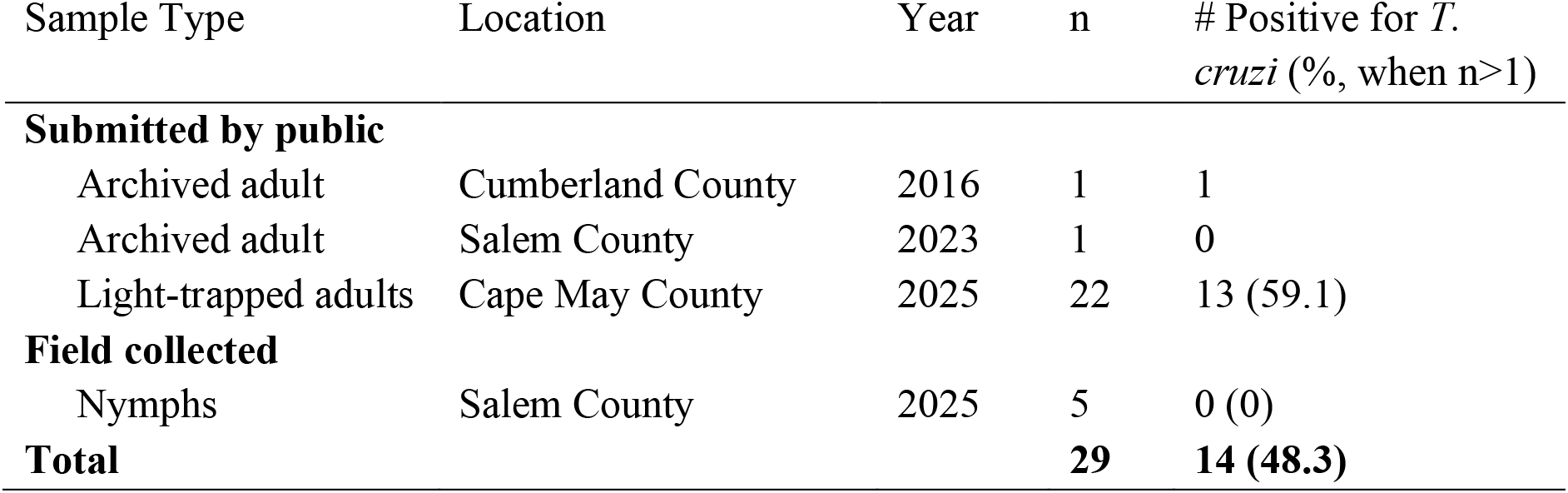
*Trypanosoma cruzi* DNA detection in *Triatoma sanguisuga* gut extractions, southern New Jersey, 2016–2025.

## Discussion

We report the first verified field collections of *Triatoma sanguisuga* in New Jersey and the first detection of *Trypanosoma cruzi* in triatomines from the state. *Triatoma sanguisuga* was collected at multiple sites in southern New Jersey, whereas neither triatomines nor *Trypanosoma cruzi* were detected at the central New Jersey sites examined. At one of the New Brunswick sites, however, habitat surveys yielded the flat bug *Mezira subsetosa* (Hemiptera: Aradidae), which to our knowledge represents the first state record of this species. *Mezira subsetosa* was first reported in North America from Maryland in 2010 and has since been documented from the eastern United States from New York to Georgia (Hoebeke et al. 2019), indicating that our sampling was sufficient to recover other cryptic, bark-associated heteropterans. However, because we did not develop a systematic statewide survey, non-detection of *T. sanguisuga* outside southern New Jersey should not be interpreted as evidence of absence. Rather, these collections establish a baseline for evaluating the distribution of *T. sanguisuga* and *Trypanosoma cruzi* in the state.

Several observations support the presence of established *T. sanguisuga* populations in southern New Jersey. Community-science observations and two specimens submitted by New Jersey residents indicate that the species has likely been present in the state since at least 2016 (GBIF.org 2026), although the provenance of such records cannot always be independently verified. More importantly, field collections yielded multiple individuals and multiple life stages. The occurrence of nymphs is particularly informative because immature triatomines lack functional wings and have more limited dispersal capacity than adults (Bremond et al. 2014).

Collectively, these observations are more consistent with local populations than with isolated introductions, although systematic sampling will be required to determine their geographic extent and persistence.

Phylogenetic analysis of *cox1* sequences suggested possible geographic structuring among US *T. sanguisuga* populations, with sequences from northeastern, midwestern, and southeastern regions showing some separation. However, placement of New Jersey triatomine sequences into a broader phylogenetic context was limited by the small number and restricted geographic distribution of publicly available sequences. This is particularly relevant given previously reported mitochondrial divergence within *T. sanguisuga*, which may reflect geographically differentiated populations or cryptic species (Peterson et al. 2025). Expanded sampling and additional mitochondrial and nuclear sequence data are needed to evaluate population structure across the northeastern United States.

*Trypanosoma cruzi* was detected in field-collected adult *T*

*sanguisuga* from southern New Jersey, indicating that the parasite occurs within local vector populations. In contrast, no *Trypanosoma cruzi* was detected in archived mammal samples from Middlesex County in central New Jersey, approximately 130 km north of the southern collection sites. Several sampled species, including *Procyon lotor* and *Sciurus carolinensis*, are recognized hosts of *T. sanguisuga* and have been reported infected with *Trypanosoma cruzi* (Balasubramanian et al. 2022, Busselman and Hamer 2022, Torhorst et al. 2023). However, these mammal samples were opportunistically available from a previous study rather than collected as part of a spatially matched reservoir survey, and the negative results therefore should not be interpreted as evidence that *Trypanosoma cruzi* is absent from central New Jersey. Geographic variation in vector abundance, reservoir infection, and parasite transmission could all contribute to the contrasting results between southern and central New Jersey.

The proportion of adult *T. sanguisuga* testing positive for *Trypanosoma cruzi* was broadly comparable to infection prevalences reported elsewhere in the United States, where rates of approximately 45–65% have frequently been observed (Cesa et al. 2011, Curtis-Robles et al. 2018, Dumonteil et al. 2024). Data from the northeastern United States remain limited, although infected *T. sanguisuga* have been reported from Delaware and Virginia (Curtis-Robles et al. 2018, Peterson et al. 2024). We detected *Trypanosoma cruzi* DNA in adults from Cape May County but not in nymphs from Salem County. Given the small sample sizes and different collection locations, this contrast should be interpreted cautiously. However, previous studies have reported much lower *Trypanosoma cruzi* infection prevalence in nymphs than adults (Curtis-Robles et al. 2018). This pattern may reflect greater cumulative exposure among adults through repeated blood feeding, as well as ontogenetic differences in host use that alter exposure to vertebrate hosts with differing reservoir competence or infection prevalence (Waleckx et al. 2014, Beatty et al. 2025).

Even though *Trypanosoma cruzi* only occurs in the gut and Malpighian tubules of triatomines (Schaub 2021), *Trypanosoma cruzi* DNA was detected in gut extractions as well as leg extractions from adult *T. sanguisuga*. Because live specimens were held together in shared containers before processing, cross-contamination among bugs remains a plausible explanation, particularly given that *Trypanosoma cruzi* DNA can persist in dried feces for months (Hamano et al. 2001). Accordingly, gut-positive/leg-negative specimens provide the strongest evidence of internal infection, whereas specimens with both gut and leg positivity should be interpreted more cautiously (Hamano et al. 2001, Alavez-Rosas et al. 2024). We explored multiple surface sterilization protocols, but we found that putative contamination on the legs was not adequately resolved. Future investigation of surface sterilization protocols and the use of alternative tissues as negative controls should be considered to limit false positive results.

The biology of *T. sanguisuga* is consistent with the potential for local *Trypanosoma cruzi* transmission as this species feeds on wildlife, domestic animals, and humans and is frequently encountered in and around residences (Moudy et al. 2014, Dumonteil et al. 2024, Peterson et al. 2024). However, the presence of infected vectors does not establish the likelihood or efficiency of transmission to humans. *Triatoma sanguisuga* generally exhibits longer post-feeding defecation intervals than some more efficient *Trypanosoma cruzi* vectors, potentially reducing opportunities for transmission to humans through contact with infectious feces (Killets et al. 2025). In contrast, delayed defecation may be less consequential for sylvatic transmission cycles, as wildlife can also acquire *Trypanosoma cruzi* through oral routes, including the ingestion of infected vectors or prey (Jansen et al. 2017). Thus, although infected *T. sanguisuga* populations may contribute to local parasite circulation, their epidemiological significance remains uncertain. Further investigation of local host use, vector abundance, seasonal activity, and infection prevalence will be essential for assessing their contributions to sylvatic transmission and potential human exposure.

This study combined targeted field collections with homeowner submissions, community-science records, and archived mammal samples to provide an initial characterization of *T. sanguisuga* and *Trypanosoma cruzi* in New Jersey. Although the sampling design does not permit estimation of statewide distribution, abundance, or infection prevalence, the findings establish that *T. sanguisuga* occurs in southern New Jersey and that *Trypanosoma cruzi* is present in at least some local triatomines. Systematic surveys across the state, coupled with reservoir sampling, blood-meal analysis, and expanded genetic characterization will be necessary to define the distribution, ecology, and population structure of *T. sanguisuga* at the northeastern extent of its range. While more research is needed to fully appreciate the epidemiologic significance of these findings, this report provides healthcare providers and public health practitioners preliminary information about the presence of this pathogen and its vector in New Jersey.

## Data Availability

All data produced in the present study are available upon reasonable request to the authors. Sequences will be made available on GenBank.

## Acknowledgements

We are grateful to Joseph Ingerson-Mahar and Salvatore Mangiafico of Rutgers Cooperative Extension for generously sharing their expertise in entomology and for their knowledge, guidance, and dedication to public outreach. We would also like to thank Hanna Cortes, Hunt Kinnaird, and Grace Ferry for triatomine collection assistance as well as a resident of Cape May County for providing us with adult kissing bugs caught in light traps. Finally, we would like to acknowledge Gabriel Hamer and Sarah Hamer for providing reagents for preliminary pathogen screening and Francisco Ferreira for the suggestion to screen mammal samples for *Trypanosoma cruzi*.

## CRediT Author Contribution Statement

HEH: Conceptualization, Data curation, Formal Analysis, Investigation, Methodology, Project administration, Visualization, Writing – original draft, Writing – review & editing; ARK: Investigation, Methodology; JKP: Funding acquisition, Investigation, Methodology, Resources, Writing – review & editing; DMF: Conceptualization, Funding acquisition, Investigation, Methodology, Resources, Supervision, Writing – review & editing.

